# The Vanishing White Matter registry: a unique source of natural history data

**DOI:** 10.64898/2026.09.21.26363197

**Authors:** Merel C. Postema, Romy J. van Voorst, Eline M.C. Hamilton, Daphne H. Schoenmakers, Menno D. Stellingwerff, Marjan E. Steenweg, Renate J. Verbeek, Marjo S. van der Knaap

## Abstract

The Vanishing White Matter (VWM) Registry is a single-center database containing longitudinal survey data from >470 untreated patients with DNA- and MRI-confirmed VWM. VWM is a rare genetic disorder of the brain’s white matter with an estimated live birth incidence of 1 per 100,000 and prevalence of 1.3 per 1,000,000 people in the Netherlands. The Registry represents a remarkably extensive collection of systematically gathered information on this disease, enabling crucial research into the natural history of VWM and providing a valuable resource for control arms in clinical trials. Data, initially collected on paper from 2004, have been maintained electronically since 2018, and include validated scales alongside a disease-specific customized questionnaire covering clinical, functional, and psychosocial domains. Survey packages, available in eight languages, are sent one to four times a year, depending on patient age. Built-in validity checks and review by pediatric and adult neurologists ensure data quality. The dataset is continuously updated with questionnaires expanding its scope, and structured for analysis and reuse in research on VWM.

## Background & Summary

The Vanishing White Matter (VWM) Registry is an ongoing, single-center registry coordinated by the Amsterdam Leukodystrophy Center at Amsterdam UMC. At the time of its establishment in January 2004, VWM had only recently been recognized as a distinct disorder (Leegwater, Vermeulen, Konst, et al., 2001). Very little was known about the disease, including its natural history, clinical progression, and variability. The creation of a systematic, longitudinal registry was therefore intended to provide a valuable resource for future research, laying the foundation for understanding VWM and supporting the design of clinical studies (Jonker, Bakker, Kurz, & Plueschke, 2022). Since 2018, all data have been collected electronically using Castor EDC.

The registry database is regularly updated to adapt to evolving needs, ensuring its sustainability and quality (Plueschke et al., 2025). For example, recent addition of new instruments supports the registry’s sustainability, with stakeholder communication identifying areas for further enhancement. The registry’s quality is upheld through the use of performance indicators, such as the EMA guideline on registry-based studies (European Medicines Agency, 2021), to guide updates (Jonker et al., 2026).

VWM is a rare genetic disorder with an estimated live birth incidence of 1 per 100,000 and prevalence of 1.3 per 1,000,000 individuals in the Netherlands (Hamilton et al., 2018). It is one of the more prevalent leukodystrophies, that is, a group of disorders featured by genetically determined abnormality of the brain’s white matter. VWM is caused by biallelic pathogenic variants in any of the five genes encoding the eukaryotic translation initiation factor 2B (eIF2B): *EIF2B1–EIF2B5*. The disease can manifest at any age, and earlier disease onset is typically linked to a more rapid and severe clinical course and earlier death (van der Knaap, Pronk, & Scheper, 2006).

Data on VWM have been collected through several initiatives, including the Myelin Disorders Biorepository Project (Children’s Hospital of Philadelphia) and the VWM Registry from the Amsterdam Leukodystrophy Center at Amsterdam UMC, which is the focus of this paper. The VWM registry collects longitudinal data from patients across different ages and countries, providing a rich resource to study the natural history and clinical variability of VWM. The dataset has contributed to several publications.

For example, Hamilton et al. used longitudinal registry data up to October 2016, including 296 patients, to investigate disease progression using the Health Utilities Index (HUI) and the Guy’s Neurological Disability Scale (GNDS) (Hamilton et al., 2018). Their analysis confirmed that age of onset strongly predicts disease course: early-onset patients have a faster decline and higher mortality. Additionally, the age of onset predicts the disease phenotype: childhood onset more commonly presents with motor problems, whereas cognitive problems are more frequent in adolescent and adult-onset cases. Patients with onset at or after four years generally experience a milder disease progression compared to patients with onset before four years (Hamilton et al., 2018).

Furthermore, Schoenmakers et al. conducted a cross-sectional study to evaluate the results and usability of the Vineland Adaptive Behavior Scales, Third Edition (Vineland-3) as an outcome measure for VWM patients. This study included 64 participants, of which 42 were recruited through the VWM Registry between May 2021 and July 2022. They concluded that the Vineland-3 is a useful tool for assessing the cognitive, behavioral, and psychiatric difficulties that affect daily life in VWM patients (Schoenmakers et al., 2024).

Stellingwerff et al. investigated whether quantitative MRI could detect characteristic brain changes in VWM and metachromatic leukodystrophy (MLD) and reflect clinical severity. The study included 37 VWM patients from the VWM Registry, enrolled between 2020 and 2023, with 3 Tesla MRI data and clinical assessments, including the HUI, Vineland-3 and the Gross Motor Function Classification for Metachromatic Leukodystrophy (GMFC-MLD). Of the quantitative MRI measures, the myelin water fraction showed the strongest correlations with clinical measures in both VWM and MLD patients (Stellingwerff et al., 2025).

Van Voorst et al. conducted a cross-sectional study on the impact of VWM on 100 unaffected family members of VWM patients, recruited via the VWM Registry. Data were collected using standardized questionnaires assessing health-related quality of life and psychosocial functioning. The study found that VWM impacted all family members, particularly mothers and partners of VWM patients, who reported reduced quality of life (van Voorst, Schoenmakers, van Beelen, et al., 2025). The main drivers for this were identified.

The VWM registry includes a comprehensive set of longitudinal clinical, functional, and psychosocial data, including a disease-specific customized questionnaire and validated scales. These data can support research on the natural history and progression of VWM, development of clinical outcome measures, genotype–phenotype correlations, and serve as a reference for clinical trial design (Schoenmakers et al., 2023; van der Knaap et al., 2022). In these papers related to trial design, VWM registry data were utilized to conduct power calculations (Schoenmakers et al., 2023) and to characterize disease progression across different age groups (van der Knaap et al., 2022).

## Methods

### Data Collection and Management

The VWM Registry collects longitudinal survey data from patients with DNA- and MRI-confirmed VWM across the world, predominantly from Europe (58%), North-America (18%) and South America (10%) (based on data export from 03-JUN-2026). Prospective data collection began in 2004 on paper and, since 2018, has been fully electronic via Castor EDC. Genetic conformation became available in 2001 and 2002 (Leegwater, Vermeulen, Könst, et al., 2001; van der Knaap et al., 2002). To ensure data completeness, we also entered retrospective data from available clinical records and from the self-developed Clinical History (CH) questionnaire, customized to VWM. Participants were recruited through two primary channels: (1) families who directly contacted the research team expressing their interest in participating, and (2) individuals who were referred by their physicians. To date, the VWM Registry offers survey packages in eight languages: English (US), Dutch (Netherlands), German (Standard), French (European), Italian, Polish, Portuguese (European), and Spanish (European). In certain cases, different variants of the same language are used interchangeably within the same survey package. For example, the official translation of Health Utilities Preschool (HuPS) is available in Brazilian Portuguese, while other surveys within the Portuguese survey package are in European Portuguese. However, within each survey, the language variant remains consistent.

### Ethics approval, consent and eligibility criteria

The study was approved by the medical ethical committee (Medisch Ethische Toetsingscommissie VU medical center, reference numbers 2004-03/194 and 2018.588).

To be eligible, participants must meet two criteria: (1) signed informed consent, and (2) confirmed VWM, based on both genetic results and MRI findings (van Voorst, Schoenmakers, Bonkowsky, et al., 2025). Before enrolling in the VWM Registry, patients with VWM or their guardians provide informed consent, which allows the collection of clinical records, MRI scans, and genetic test results. Additionally, assent is obtained from individuals aged 12 -16, and from those aged 16 and older who are not mentally competent to provide informed consent.

In 2023, participants of the VWM registry were asked via email to renew their consent for sharing their encoded clinical data with external parties, including physicians abroad, researchers inside and outside the EU, and companies or (semi)governmental bodies involved in VWM treatment development and healthcare policy. Nonresponse after three reminders was considered implicit consent.

### Experimental design

Most participants never visit the Amsterdam UMC and solely contribute by completing surveys online. After enrollment, they receive email invitations to fill out questionnaires and scoring forms, which are stored directly in the database. Surveys can also be completed by the parents, another family member, guardian, or physician. Medical records are also collected and reviewed to gain a more comprehensive understanding of the patient’s condition.

The frequency of the survey invitations depends on the age of the participant: four times per year for children aged 0–<4, twice per year for those aged 4–<8, and once per year for participants aged 8 years and older. Each round includes up to four questionnaires and six score forms, together taking about 30 minutes to complete.

Participants who visit the Amsterdam UMC for standard patient care may additionally contribute data from physical and neurological examinations. During these visits, the Vineland-3 and the Modified Rankin Scale can also be administered. These measures can alternatively be collected via video consultation.

### Clinical and Functional Assessments: core data set

The registry includes validated scales, including the HUI (i.e., HUI Mark 3 proxy version covering vision, hearing, speech, ambulation, dexterity, emotion, cognition and pain, as well as one HUI Mark 2 item covering self-care) (W. J. Furlong, Feeny, Torrance, & Barr, 2001), GNDS (Fraser & McGurl, 2007), Gross Motor Function Classification System – Extended & Revised (GMFCS-E&R) (Piscitelli, Ferrarello, Ugolini, Verola, & Pellicciari, 2021), Communication Function Classification System (CFCS) (Hidecker et al., 2011), and Manual Ability Classification System (MACS) (Eliasson et al., 2006), alongside the customized CH questionnaire, which contains a rich set of questions for first inventory (all questions) and follow-up (subset of questions). The CH questions that assess the age of disease onset, the age at which participants lose the ability to walk without and with support, as well as instances of rapid deterioration, and age at death, are considered core data elements.

Later additions include the EuroQol 5-Dimension 5-Level (EQ5D-5L) (Herdman et al., 2011), EuroQol 5-Dimension 3-Level Youth (EQ5D-3Y) caregiver version (Rabin & de Charro, 2001), Expressive Language Function Classification in Metachromatic Leukodystrophy (ELFC-MLD) (Boucher et al., 2015; Kehrer et al., 2014), Eating and Drinking Ability Classification System (EDACS) (Bykova, Frank, & Girolami, 2023), Gross Motor Function Classification-Metachromatic Leukodystrophy (GMFC-MLD) (Gavazzi et al., 2023), HuPS (W. Furlong et al., 2023; Viani et al., 2024) and mini-MACS (Eliasson, Ullenhag, Wahlström, & Krumlinde-Sundholm, 2017).

Table 1 shows which instruments are sent for each age group. Where possible, validated translations are used. If an official translation of an instrument is not available, it is either excluded from the survey packages or, in cases like the CH questionnaire, translated by leukodystrophy clinical experts in the field, being native speakers.

**Table 1.** Instruments collected via surveys in the VWM Registry.

| Age category | Instrument(s)<sup>1</sup> |  |  |  |  |  |  |  |  |  |  |  |  |
| --- | --- | --- | --- | --- | --- | --- | --- | --- | --- | --- | --- | --- | --- |
|  | 1 | 2 | 3 | 4a | 4b | 5 | 6 | 7 | 8 | 9a | 9b | 10a | 10b |
| 0 to < 2 years | X |  |  |  | X |  | X |  |  |  |  |  | X |
| 2 to < 4 years | X | X | X |  | X | X | X | X |  | (X) | X |  | X |
| 4 to < 8 years | X | X | X |  | X | X | X | X |  | X |  | X |  |
| 8 to < 18 years | X | X | X |  | X | X | X | X | X | X |  | X |  |
| 18 years and older | X | X | X | X |  | X | X | X | X | X |  | X |  |

<sup>1</sup> **1= CH questionnaire and clinical records.** Data collected since 25-MAY-1988. Available in all eight languages; **2 = CFCS.** Data collected since 23-NOV-2009. Available in all eight languages; **3 = EDACS.** Data collected since 29-JAN-2025. Available in Dutch and English; **4a = EQ5D-5L.** Data collected since 11-MAR-2026. Available for Dutch, English, German, French and Spanish; **4b = EQ5D-3Y** caregiver version. Data collected since 11-MAR-2026. Available for Dutch, English, German, French and Spanish; **5 = ELFC-MLD.** Data collected since 29-JAN- 2025, and currently only available in Dutch and English; **6 = GMFCS-E&R.** Data collected since 23-NOV-2009. Available in all eight languages; **7 = GMFC-MLD.** Data collected since 29-JAN- 2025, and currently only available in Dutch and English; **8 = GNDS.** Data collected since 16-JAN-2004. Available in all eight languages; **9a = HUI 2&3.** Data collected since 16-JAN-2004. Since 11-MAR-2026 only collected from 4 years and older. For age category 2 to < 4 years replaced by instrument 9b (HuPS), hence the X in parentheses. Available in all eight languages; **9b = HuPS.** Data collected since 11-MAR-2026 for ages 2 to < 4 years. Available in Dutch, English and Brazilian Portuguese; **10a = MACS.** Data collected since 23-NOV-2009. Available in all eight languages; **10b = Mini-MACS.** Data collected since 3-OCT-2025 for all eight languages except German, French and Polish.

### Clinical and Functional Assessments: optional data set

The optional dataset (not listed in Table 1) contains data gathered outside the standard survey packages, including the institute for Medical Technology Assessment (iMTA) Productivity Cost Questionnaire (iPCQ) (C. Bouwmans et al., 2015), the iMTA Medical Consumption Questionnaire (iMCQ) (C Bouwmans et al., 2013), and the EuroQol 5-Dimension 3-Level Youth (EQ5D-3Y) self-report version (Rabin & de Charro, 2001). Additionally, the Modified Rankin Scale (mRS) (Banks & Marotta, 2007) and the Vineland-3 (Sparrow, Cicchetti, & Saulnier, 2016) are optional and can be administered during Amsterdam UMC visits. Also, data elements from the CH questionnaire that do not capture core disease outcomes, like information about consanguinity, pregnancy, neonatal period, early cognitive development, speech development, and implementation of preventive measures, are considered optional.

### Computational processing

Data are collected using Castor EDC and exported in CSV format. R scripts developed to merge the raw data into a single analysis-ready table can be provided once the data becomes accessible.

## Data Record

The VWM registry dataset is hosted on Castor EDC, with data stored on a server located in the Netherlands. The data can be exported in CSV or SPSS formats. We work with CSV format for analysis, as it is the format used in the associated R scripts. The core dataset includes the following data elements, in accordance with the EMA guideline on registry-based studies (European Medicines Agency, 2021):

- **Administrative Information**: The dataset includes the center’s name (i.e., Amsterdam UMC), the informed consent date and, where applicable, details such as the reason for early termination and the date of the last participant contact. These data points are recorded in the electronic Case Report Form (eCRF). The participant creation date, which matches the registry entry date, is also available in Castor EDC.
- **Patient Data**: The eCRF collects the participant’s month and year of birth, as well as biological sex and country of residence.
- **Disease**: All participants have a DNA- and MRI-confirmed diagnosis of VWM. The eCRF captures the specific genetic mutations identified in each participant. While MRI images are stored separately and are not systematically collected after entering the registry, the date of the MRI and the participant’s age at the time of the MRI are recorded during clinical visits. We do not collect the exact date of diagnosis, as it is influenced by factors unrelated to the disease itself and can vary significantly from the date when the first clinical symptoms appear. Hence we use the latter, as reported by participants in a survey, to indicate the start of the disease. Disease severity is assessed indirectly based on responses to survey questions that gauge functional abilities and limitations. Core disease outcomes include the age at which participants lose the ability to walk without and with support, instances of rapid deterioration, and age at death.
- **Co-morbidities**: Co-morbidities present during the screening phase are recorded in the eCRF and can be further specified in a text field. Although past comorbidities are not explicitly queried, they may be noted in the text field as well.
- **Disease-targeting treatments**: As a rule, participants are non-medicated for the disease, which facilitates the study of the disease’s natural progression. Patients who enter a clinical trial may remain part of the registry, but only data up till trial start is used for analyses and data sharing. The eCRF captures information about trial participation, including trial name and start date.
- **Relevant concomitant therapies**: Information on other medications for neurological symptoms related to VWM or for unrelated diseases, including dosage, frequency, and start and stop dates, can be recorded in the eCRF, but there is no requirement for their regular documentation.
- **Adverse events**: Not applicable (see above).
- **Current pregnancy**: This information is not collected, as VWM is primarily a childhood disorder. Female patients often have ovarian failure and pregnancies after diagnosis are exceptional (Fogli et al., 2003).
- **Patient-reported outcomes in clinical practice**: Validated instruments, as well as the customized CH questionnaire, are distributed to participants via survey packages, as listed in Table 1.

## Data Overview

Figure 1 illustrates the growth of the VWM Registry over time, showing for the standard packages the number of responses per survey for each year.

**Figure 1.**
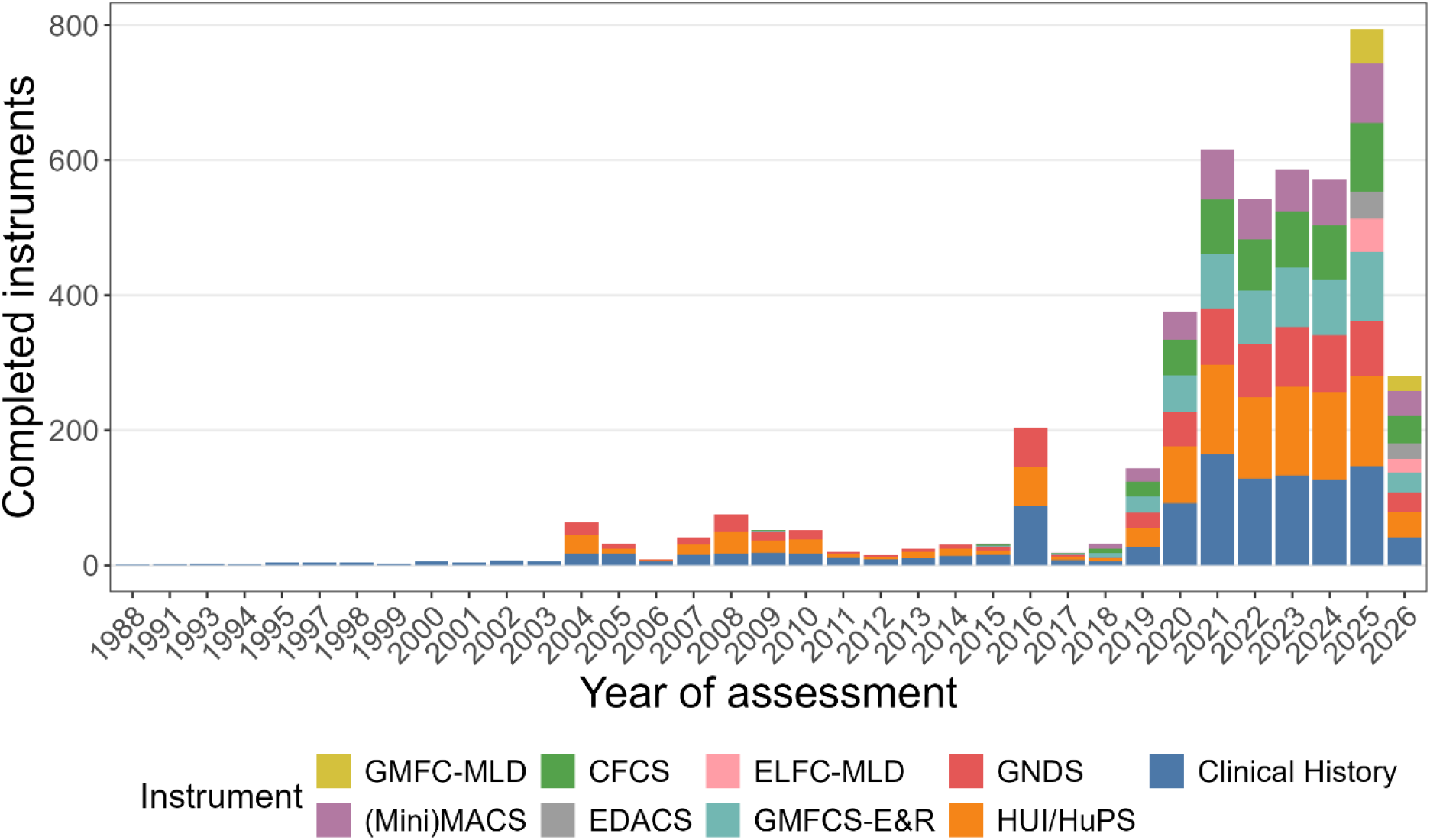
An overview of the number of different instruments completed by participants in the VWM registry over time (based on data export from 03-JUN-2026). Notably, while prospective data collection for the registry started in 2004, a few retrospectively completed CH questionnaires were available and entered to ensure inclusion for analyses.

## Technical Validation

The HMA-EMA Data Quality Framework (DQF) (HMA-EMA, 2023) distinguishes three categories of data quality elements, referred to as “determinants”: foundational, intrinsic, and question-specific determinants. Each determinant is described below and, where possible, linked to the four main components of data quality: consistency, completeness, accuracy, and representativeness (European Medicines Agency, 2021). In addition, the risk-based assessment procedure and compliance framework for the VWM registry data are discussed.

### Foundational determinants

Data are electronically collected via Castor EDC/CDMS, which is certified for ISO 27001, ISO 27002, and NEN7510. The system incorporates an integrated standard audit trail to ensure data quality throughout the collection process. In addition, robust security measures are implemented, with user-specific access rights assigned according to roles, and regular data backups conducted to further safeguard data integrity. Data processing steps are well documented through R scripts with version control. Consistency is ensured, among others, by clearly defining measurement units and providing additional help text where necessary.

Recently, consistency across translations was validated for key items by back-translating the questions into English and comparing them. Whenever possible, official translations were used (e.g., for standard instruments), and other translations were provided by leukodystrophy clinical experts, who are native speakers. While a standard terminology is not yet in place, efforts are ongoing to enhance the database and incorporate Clinical Data Interchange Standards Consortium (CDISC) Standards by mapping the data to a Study Data Tabulation Model (SDTM) and an Analysis Data Model (ADaM) (Consortium, 2026). In order to improve accuracy and reliability for key disease progression variables - specifically, loss of walking without support, loss of walking with support, episodes of rapid deterioration, and age at death - these variables are collected both via the CH questionnaire and study/eCRF data. As survey responses are sometimes inconsistent over time, curated study data are used as the definitive source for these outcomes. Finally, all changes to the database are carefully documented, with each version archived in DANS’s preferred format and verified with checksums to ensure data integrity.

### Intrinsic determinants

Castor EDC includes several built-in quality checks that trigger warnings when a participant enters an unexpected response. These checks are applied within individual questionnaires, including HUI, GNDS, and the CH questionnaire, as well as across different instruments, including the aforementioned ones as well as EDACS, ELFC-MLD and GMFC-MLD. Table 2 provides an overview of these checks, detailing the items involved and the expected consistency rules. These built-in validations help ensure that the collected data are internally consistent and reduce errors at the point of entry. Furthermore, range constraints were added for numeric variables, when possible.

**Table 2.** Built-in quality checks in Castor EDC across and within instruments.

| Check | Instrument(s) |  |  |  |  |  |  |  |  |  | Description |
| --- | --- | --- | --- | --- | --- | --- | --- | --- | --- | --- | --- |
|  | 1 | 2 | 3 | 4 | 5 | 6 | 7 | 8 | 9 | 10 |  |
| Speech/<br>communication |  | X |  |  | X |  |  |  |  |  | Communication functioning should correspond to expressive language functioning |
|  |  | X |  |  |  |  |  |  | X |  | Communication functioning should correspond to speech abilities (HUI05) |
|  |  |  |  |  |  |  |  | X | X |  | Speech abilities (HUI05) should correspond to GNDS speech/communication ratings |
|  |  |  |  |  |  |  |  |  | X |  | Speech with unfamiliar (HUI05) vs. familiar people (HUI06) should correspond. |
| Mobility |  |  |  |  |  | X | X |  |  |  | Gross motor should correspond to motor functioning |
|  |  |  |  |  |  |  | X | X |  |  | Gross motor should correspond to GNDS lower limb disability |
|  |  |  |  |  |  |  | X |  | X |  | Gross motor should correspond to ambulation score (HUI09) |
|  | X |  |  |  |  |  | X |  |  |  | Gross motor should correspond to wheelchair dependency |
|  |  |  |  |  |  | X |  |  | X |  | Motor functioning should correspond to ambulation score (HUI09) |
|  |  |  |  |  |  |  |  | X |  |  | GNDS indoor and outdoor movement items should match |
|  |  |  |  |  |  |  |  | X | X |  | GNDS lower limb disability should correspond to ambulation score (HUI09) |
|  | X |  |  |  |  |  |  |  | X |  | Ambulation score (HUI09) should match wheelchair use |
|  | X |  |  |  |  |  |  |  |  |  | Wheelchair dependency should match with loss of walking without support |
| Fine Motor<br>functioning |  |  |  |  |  |  |  |  | X | X | Manual dexterity should match hand/finger function (HUI10) |
|  |  |  |  |  |  |  |  | X | X |  | GNDS upper limb disability should correspond to hand/finger function(HUI10) |
|  |  |  |  |  |  |  |  |  | X |  | Hand/finger usage (HUI10) should correspond to self-care level (HUI13) |
| Emotions |  |  |  |  |  |  |  | X | X |  | GNDS mood assessment should correspond to usual feelings (HUI07) |
|  |  |  |  |  |  |  |  |  | X |  | Usual feelings (HUI07) should correspond to overall feelings (HUI14) |
| Cognition |  |  |  |  |  |  |  | X | X |  | GNDS cognitive ability should correspond to thinking and problem solving level (HUI12) |
|  | X |  |  |  |  |  |  |  |  |  | Current intelligence should be equal to or lower than maximum intelligence |
| Pain |  |  |  |  |  |  |  |  | X |  | Usual pain level (HUI08) should match general pain level (HUI15) |
| Eating and<br>drinking |  |  | X |  |  |  |  | X |  |  | Eating and drink ability should match GNDS swallowing ability |
| Vision |  |  |  |  |  |  |  | X | X |  | GNDS visual ability should correspond to near (HUI01) and far (HUI02) vision score. |

<sup>1</sup> 1= Clinical History questionnaire, 2 = CFCS, 3 = EDACS, 4 = EQ5D-5(L/3Y); note that there are no quality checks for this survey, it is only listed for completeness, 5 = ELFC-MLD, 6 = GMFCS-E&R, 7 = GMFC-MLD, 8 = GNDS, 9 =HUI / HuPS, 10 = (Mini) MACS.

Plausibility is periodically assessed through automated R scripts that track changes in scores over time, including GMFC-MLD, HUI, HuPS and (Mini) MACS. Any differences greater than two points compared to the previous submission are flagged for review by pediatric and adult neurologists, and corrected if needed (based on four-eye verification principle) to maintain data quality. Data accuracy is improved through outlier detection, source data verification and monitoring. Precision is further enhanced by reporting age in years and months, and by using age-specific surveys and survey packages.

### Question specific determinants

Regular checks for data completeness are conducted using automated R scripts and the “naniar” package to visualize missing values (Tierney & Cook, 2023). The representativeness of the data cannot be assessed against other datasets, as the VWM registry is by far the largest dataset on VWM available, and other data are limited.

### Risk-based assessment

Team Quality Assurance Clinical Research (Amsterdam UMC) has developed a guidance document on quality control of non-WMO (i.e., non-Medical Research Involving Human Subjects Act) data registries that do not undergo clinical monitoring audits by the Clinical Monitoring Center. The VWM Registry follows this guidance, which is based, among other sources, on the EMA guidelines for registry-based studies (European Medicines Agency, 2021). The recommended checks are applied at least annually to maintain data quality. A risk-based sampling approach is used, were data quality is evaluated across five pillars: consistency, completeness, accuracy, timeliness and traceability. For risk classification, three categories of data points are distinguished: 1) critical data points, which are essential for the primary objectives of the registry, 2) important data points, which are relevant for certain stakeholders, but not essential for all, and 3) supportive data points, such as exploratory variables or free-text fields, which are excluded from sampling.

### Compliance framework

The data quality processes outlined above are documented in a standard operating procedure (SOP), which enhances consistency. In compliance with Good Clinical Practice (GCP) guidelines, all information related to the VWM registry is systematically organized within a Trial Master File (TMF). All team members receive the necessary training, with their certification records stored in the TMF. These processes can be integrated within the framework of a standard Quality Management System (QMS), which includes features such as version control, audit trails, document ownership, and periodic document reviews (NFU, 2023).

## Data Availability

The metadata of the VWM registry dataset are available in the European Directory of Registries (ERDRI) as well as the Amsterdam UMC DataverseNL repository (doi:10.34894/LS1ZGD). The VWM registry is also findable on Orphanet (ORPHA:135).
The pseudonymised dataset, containing selected variables from merged survey responses in eight languages along with curated clinical data, can be made accessible upon request.

https://dataverse.nl/dataset.xhtml?persistentId=doi:10.34894/LS1ZGD

https://eu-rd-platform.jrc.ec.europa.eu/mdr/detail/VWM-Registry

## Usage Notes

Secondary data users interested in both the core and optional datasets should be aware that the same EQ5D-5L survey is used in different contexts. Specifically, in the standard survey package, which comprises the core dataset, the EQ5D-5L is used to gather data about the patient. In contrast, in the non-standard survey package, which constitutes the optional dataset, the same survey is used to gather data about the caregiver. Automated R scripts, run after data export, differentiate between the core and optional EQ5D-5L data. It is essential to maintain this distinction during analysis to ensure data quality.

## Data Availability

The metadata of the VWM registry dataset are available in the European Directory of Registries (ERDRI) as well as the Amsterdam UMC DataverseNL repository (doi:10.34894/LS1ZGD). The VWM registry is also findable on Orphanet (ORPHA:135).

## Data Sharing and Governance

The pseudonymised dataset, containing selected variables from merged survey responses in eight languages along with curated clinical data, can be made accessible upon request. Only the data relevant to answer the study question will be shared, in accordance with a data usage agreement developed in consultation with the Legal Research Support department of Amsterdam UMC. Highly sensitive data or potentially identifiable data, including genetic mutations, month and year of birth, and country of residence, will not be shared. Requests for data access must include a clear description of the intended use, as well as a specified end date for sharing (typically 16 months). Fees may apply for commercial or profit-driven purposes.

Each data request is assessed by the data access committee advised by the VWM consortium, which is an international academic network of neurologists specialized in leukodystrophies, and the VWM Families Foundation, which is a patient advocacy group. The VWM Consortium and VWM Families Foundation advise Amsterdam UMC on decisions, while Amsterdam UMC remains end responsible. Data can be released on an Azure-based digital research environment (myDRE) platform, where data can be accessed, analyzed and only downloaded in aggregated form after permission of the host. In this way the data are released in a secure way with maximum privacy protection.

## Code Availability

Custom R scripts have been used for data pre-processing, including merging survey responses across languages, harmonizing checkbox variables by systematically renaming them with numeric suffixes, and combining survey and study data into a single dataset for analysis. Data pre-processing has been performed in R (version 4.6.0) (R Core Team, 2025). All custom scripts will be provided once data are made accessible.

## Acknowledgements, Author Contributions & Competing Interests

*Acknowledgements*

The authors would like to thank all VWM patients and their family members for their contributions to the Vanishing White Matter Registry, all colleagues for referring patients or providing information, and acknowledge the contributions of the Vanishing White Matter Consortium (https://www.vwmconsortium.org/about-us/).

## Author Contributions

M.C.P. wrote the original draft and conducted data curation. R.J.v.V. created Figure 1 and R.J.V. built the Castor database. R.J.v.V., E.M.C.H., D.H.S. and M.D.S. were involved in project administration. M.S.v.d.K. supervised the project and further contributed through conceptualization and funding acquisition. All authors read and approved the final manuscript.

### Competing Interests

The authors declare that there are no direct commercial conflicts of interest related to the data presented in this paper. The VWM Registry is partially supported by funding from pharmaceutical companies. These funds are solely used for the ongoing maintenance and operation of the registry. The authors affirm that the funding structure does not influence the development, management, or availability of the dataset described.

## Notes

### Funding

- NWO, Spinozapremie: 2008

